# Genetic Factors and the Outcomes of Transplant-Associated Thrombotic Microangiopathies

**DOI:** 10.1101/2025.07.10.25331317

**Authors:** Daijing Nie, Lili Liu

## Abstract

Transplant-associated thrombotic microangiopathy (TA–TMA) is a severe endothelial complication following allogeneic hematopoietic stem-cell transplantation (allo-HSCT), associated with high mortality when not promptly diagnosed and treated. This study aimed to delineate the genetic landscape associated with TA–TMA and assess its impact on clinical outcomes. We retrospectively analyzed 1069 allo-HSCT recipients between January 2016 and May 2020, identifying 131 patients who met diagnostic criteria for TA–TMA (incidence rate: 12.25%). Genomic DNA sequencing was performed targeting 17 complement-related genes, identifying 74 genetic variants in 58 TA–TMA patients, including seven large deletions within the *CFH–CFHR5* locus. Survival analysis indicated significantly poorer outcomes for TA–TMA patients compared to non-TMA patients (24.4% vs 62.4% survival at maximum follow-up, *p* = 0.025). However, cumulative incidence curves revealed no significant difference in TA–TMA onset between genetic variant carriers and non-carriers. These findings underscore the complexity of TA–TMA pathogenesis, suggesting that genetic predisposition alone is insufficient without additional endothelial insults. The limited predictive value of individual markers highlights the need for integrated biomarker strategies. Future research should focus on refining risk stratification models incorporating comprehensive genetic profiles, dynamic biomarkers, and longitudinal clinical parameters to enable earlier identification and targeted interventions, thereby improving post-transplant survival outcomes.

## Introduction

Transplant-associated thrombotic microangiopathy (TA–TMA) is a life-threaten -ing endothelial disorder that complicates 5–40% of allogeneic haematopoietic stem-cell transplants (allo-HSCT) and carries a reported mortality of up to 80 % when diagnosis is delayed or treatment is sub-optimal [1, 2]. Clinically, TA–TMA is characterised by microangiopathic haemolytic anaemia, thrombocytopenia, acute kidney injury, and refractory hypertension; yet these “classic” signs often emerge late in the disease course, underscoring the need for earlier diagnostic markers [3]. Over the past decade, converging evidence has implicated sustained complement activation—initiated by endothelial injury, conditioning chemotherapy, calcineurin-inhibitor exposure, graft-versus-host disease (GVHD), infection, and other post-transplant insults—as the central driver of microvascular thrombosis [4, 5].

A sizeable minority of patients appear to harbor an intrinsic susceptibility to uncontrolled complement amplification. Next-generation sequencing studies have documented rare or deleterious germline variants in both alternative-pathway and terminal-pathway genes, including *CFH, CFI, C3, CFB, CD46, THBD*, and *C9* [3]. These variants mirror those observed in atypical haemolytic uraemic syndrome (aHUS) and converge on dysregulated C3/C5 convertase activity, predisposing carriers to TMA when a “second hit” damages the endothelium. However, the penetrance of individual variants is incomplete, and the extent to which specific genetic lesions modulate clinical course or therapeutic responsiveness in the HSCT setting remains poorly defined.

To date, clinical and biochemical markers routinely monitored in transplant patients, such as blood pressure, coagulation function, liver enzymes, lactate dehydrogenase (LDH), serum creatinine, proteinuria, and viral infections, are widely available. Nevertheless, their integrated value for predicting TA–TMA occurrence and outcome has yet to be systematically evaluated. Moreover, few studies have integrated genetic risk factors, dynamic biomarker trajectories, and real-world clinical endpoints in a systematically characterized cohort to guide the timely initiation of complement inhibitors (e.g. eculizumab) that have transformed outcomes in recent observational series [6].

Against this backdrop, we conducted a comprehensive analysis of allo-HSCT recipients with diverse haematological disorders to (i) delineate the landscape of TA–TMA-associated germline and somatic complement gene abnormalities; (ii) correlate these findings with longitudinal clinical features, biochemical profiles, and time-to-TMA onset; and (iii) determine the prognostic impact of specific genetic and laboratory parameters on overall survival and renal recovery. By integrating high-resolution genomic data with granular phenotypic information, our study aims to refine the mechanistic understanding of TA–TMA and to inform personalised monitoring and therapeutic strategies in the post-transplant setting.

## Patients and Methods

### Research subjects

We retrospectively collected clinical and laboratory data from patients who underwent allo-HSCT at the Hebei Yanda Lu Daopei Hospital between January 2016 and May 2020. Clinical parameters, including blood pressure, hemoglobin level, platelet count, liver enzymes, and renal function, were monitored daily starting from the first day after transplantation. Additionally, lactate dehydrogenase (LDH), schistocyte counts, and proteinuria were assessed at least twice weekly. The threshold for transfusion was set at hemoglobin levels below 7 g/dL and platelet counts below 10 × 10^9^/L. Platelet engraftment was defined as achieving a platelet count ≥20 × 10^9^/L for three consecutive days without transfusion support.

The diagnosis of TA–TMA was established based on the following criteria: (1) elevated LDH levels; (2) proteinuria (random urine protein ≥30 mg/dL); hypertension (for patients under 18 years old: blood pressure ≥95th percentile adjusted for age, gender, and weight; for patients aged 18 or older: blood pressure ≥140/90 mmHg); (4) new-onset thrombocytopenia (platelet count *<* 50 × 10^9^/L or ≥50% decrease from baseline); (5) new-onset anemia (below age-adjusted normal limits or requiring transfusion); (6) evidence of microangiopathy (presence of schistocytes on peripheral blood smear or biopsy-confirmed microangiopathy); (7) evidence of downstream complement activation (elevated plasma sC5b-9 levels); and (8) no coagulopathy with negative Coombs test. Patients were diagnosed with TA–TMA when they met at least four of the first seven criteria plus criterion number 8. The onset date of TA–TMA was defined as the first day post-transplantation on which diagnostic criteria were fulfilled.

All patients enrolled in the HSCT database and tissue repository provided informed consent. The Institutional Review Board approved the retrospective review and analysis of electronic medical records and the HSCT database.

### Genetic Variant Identification

Genomic DNA was isolated from peripheral blood samples using standard extraction methods. Sequencing and analysis were performed for 17 genes associated with thrombotic microangiopathy and complement regulation, including *CFB, CFD, CFP, CFI, CFH, CFHR1, CFHR2, CFHR3, CFHR4, CFHR5, THBD, CD46, CD59, C3, CD40L, ADAMTS13*, and *DGKE*. For patients admitted prior to February 2019, targeted amplicon sequencing was utilized. For patients admitted after February 2019, whole-exome sequencing (WES) was performed. DNA libraries were prepared according to the manufacturer’s protocol, followed by high-throughput paired-end sequencing on an Illumina sequencing platform, achieving an average depth of coverage of at least 100× per sample. Unlike Mendelian diseases or malignant disease with clear genetic predispositions, we did not employ *in silica* predictions to exclude variation with uncertain significance. All variations with the minor allele frequency *<* 1% in normal population are included for further analysis based on gnomAD database[7, 8, 9]. Besides point variations and small insertion and deletions, we also detected copy number variations (CNVs) within the *CFH–CFHR5* locus based on read-depth analysis[1]. Deletion polymorphisms were assessed at the following genomic coordinates: *CFH* (chr1:196620408–196717234), *CFHR3* (chr1:1967433 25–196763803), *CFHR1* (chr1:196788287–196801919), *CFHR4* (chr1:196818771– 196888702), *CFHR2* (chr1:196788298–196928956), and *CFHR5* (chr1:196946067– 196979404). Candidate CNVs identified by these methods were further confirmed by multiplex ligation-dependent probe amplification.

### Statistics

Patients were divided into two groups based on whether they developed TA–TMA from the time of allo-HSCT until the end of follow-up. Kaplan–Meier analysis was employed to estimate the overall survival (OS) probabilities, with comparisons conducted using the log-rank test. The end of follow-up was defined as the date of study initiation, patient death, or last patient contact (lost to follow-up). Statistical analyses were performed using Python 3.12.9. A *P* -value *<* .05 was considered statistically significant.

## Results

### Cohort characteristics

There were 1069 patients who underwent allo-HSCT during the study period and had complete medical records. The demographic and transplantation characteristics are summarized in Table 1. Except for four patients from West Asia, all other patients were of East Asian ethnicity, with a higher proportion of pediatric patients and males. Most patients underwent transplantation for malignant hematological disorders, while the remainder were diagnosed with aplastic anemia or other bone marrow failure syndromes. All patients received myeloablative conditioning regimens. Among these transplant recipients, 131 patients met the diagnostic criteria for transplant-associated thrombotic microangiopathy (TA–TMA), yielding an incidence rate of 12.25%. Most patients received transplants from related donors, and all underwent myeloablative conditioning; a subset of patients also received total body irradiation (TBI) as part of their conditioning regimen.

**Table 1.** Patient demographics and clinical characteristics.

| Characteristic | Patients with TA-TMA (n=131) | Patients without TA-TMA (n=938) |
| --- | --- | --- |
| Median age (range), years | 18 (1–62) | 17 (0–64) |
| Male/Female, n | 81/50 | 535/403 |
| Disease diagnosis, n |  |  |
| Bone marrow failure | 13 | 93 |
| MDS | 5 | 29 |
| AML | 47 | 352 |
| ALL | 59 | 420 |
| Others<sup>a</sup> | 7 | 44 |
| Donor type, n |  |  |
| Related | 124 | 883 |
| Unrelated | 7 | 55 |
| Conditioning regimen, n |  |  |
| TBI-based | 53 | 461 |
| Non-TBI-based | 68 | 477 |

<sup>a</sup>Others include NK-cell leukemia, acute mixed-cell leukemia, hemophagocytic lymphohistiocytosis, and blastic plasmacytoid dendritic cell neoplasm, among others.

The duration of follow-up ranged from day 11 to day 1452 after transplantation, with a median follow-up time of 229 days. The time to onset of TMA ranged from day 6 to day 381 post-transplantation, with a median onset at day 71.

### Genetic Variations and the Risk

Given that the association between complement regulatory pathway genes and TA-TMA remains inconclusive, we adopted a relatively broad definition for variant identification in this study. Unlike the strict pathogenic variant criteria previously established at our center, we included all variants with a minor allele frequency (MAF) of less than 1% in the general population, regardless of in silico functional prediction results. Among the 131 TA-TMA patients, we identified a total of 74 variants from 58 patients, including seven large deletions within the *CFH–CFHR5* locus. Notably, five patients carried three or more variants. It is important to note that the analysis of CNVs in the *CFH–CFHR5* locus was limited in this study, as only 46 TA-TMA patients had available WES data; thus, the seven cases of CNVs were identified exclusively among these 46 patients (Table S1).

We subsequently divided the patients into a variant carrier group and a non-carrier group. We analyzed the cumulative incidence of TA-TMA in these two groups (Fig. 1). The cumulative incidence curves demonstrated that the presence of genetic variants did not have a statistically significant effect on the time to TA-TMA onset.

**Figure 1.**
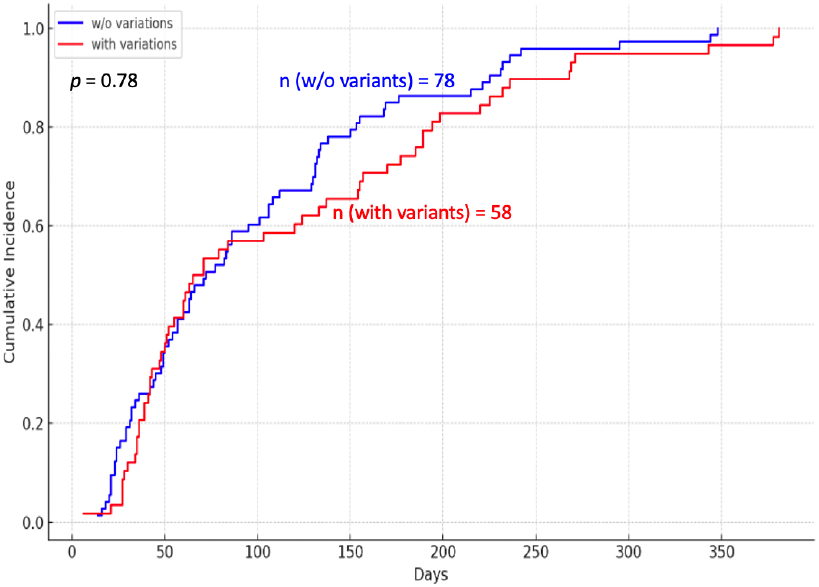
Cumulative incidence of TA-TMA in patients with and without complement-related genetic variants.

### Overall Survival

During a total follow-up period of up to 1452 days, poor outcomes were observed in 99 out of 131 patients with TMA. To further assess the impact of systemic complement activation on overall survival, we compared the prognosis of these 131 TMA patients with that of 934 allo-HSCT recipients who did not develop TMA. The results are shown in Figure 2. Although the prognosis of all-HSCT recipients without TMA was still suboptimal, their overall survival remained significantly better than that of patients who developed TMA. At the maximum follow-up of 1302 days, 586 out of 938 patients without TMA were still alive. The difference in overall survival between the TMA and non-TMA groups was statistically significant (log-rank *p* = 0.025; 62.4% vs 24.4%).

**Figure 2.**
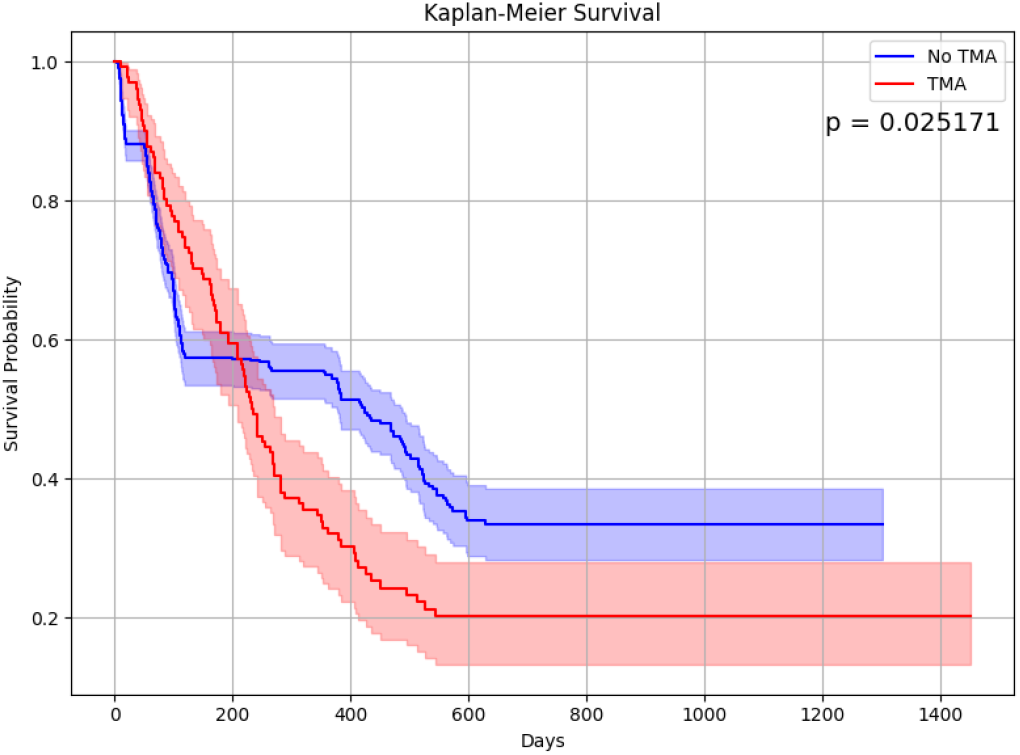
Kaplan-Meier curves of overall survival for patients with and without TMA following allogeneic HSCT.

## Discussion

In the current study, we performed an integrative analysis of genetic variations, clinical trajectories, and survival outcomes in a systematically characterized cohort of patients undergoing allo-HSCT. Our findings reinforce the notion that TA–TMA remains a significant complication following transplantation, with a markedly poorer prognosis compared to patients who did not develop the condition. The identification of complement-related genetic variants, particularly rare variants and large deletions within the *CFH–CFHR5* locus, highlights the underlying genetic susceptibility that may predispose certain individuals to dysregulated complement activation and subsequent endothelial injury.

Despite identifying some genetic variants among TA–TMA patients, we found no statistically significant difference in the cumulative incidence of TMA onset between variant carriers and non-carriers. This observation aligns with recent literature indicating that genetic predisposition alone may not be sufficient to induce TA–TMA; rather, a “second-hit” event—such as conditioning chemotherapy, calcineurin-inhibitor exposure, GVHD, or infection—appears critical in triggering overt clinical manifestations[10]. Indeed, previous studies have demonstrated that the penetrance of complement gene variants is influenced by the cumulative burden of endothelial insults and by individual variability in inflammatory responses [11, 12].

Notably, our analysis demonstrated a significant survival disadvantage for TA–TMA patients compared to non-TMA patients, underscoring the importance of early recognition and intervention. Consistent with recent observational studies, we observed that routine clinical and biochemical markers, although widely available and useful for patient monitoring, have limited predictive utility when assessed individually. This emphasizes the need for integrated, dynamic biomarker panels that incorporate genetic predisposition, complement activation status, and clinical risk factors to facilitate prompt diagnosis and targeted therapeutic decisions.

Our results also underline the emerging role of complement inhibitors such as eculizumab and ravulizumab in mitigating the progression of TA–TMA. Recent observational series suggest that timely initiation of these agents can significantly improve patient outcomes. However, the optimal timing, duration, and patient selection criteria for complement inhibitor therapy remain areas of active investigation. Our data suggest that patients harboring complement-related genetic variants, especially those with multiple pathogenic lesions or large deletions, might represent a subgroup particularly likely to benefit from prophylactic or preemptive complement blockade.

Future prospective studies are necessary to validate our findings and to refine risk stratification models by incorporating novel biomarkers, advanced genetic profiling, and longitudinal clinical assessments. Such precision medicine approaches hold promise for identifying high-risk individuals earlier in their disease trajectory, thereby enabling interventions that could substantially alter the natural history of TA–TMA and improve post-transplant survival outcomes.

## Data Availability

All data produced in the present study are available upon reasonable request to the authors.

